# Frailty in older patients with atrial fibrillation and its relationship with anticoagulant use: a multi-centred observational study in New South Wales

**DOI:** 10.1101/2024.12.20.24319406

**Authors:** Tu N Nguyen, Kenji Fujita, Sarah N Hilmer

## Abstract

**Background and aims:** Evidence of the impact of frailty on oral anticoagulant (OAC) prescription in older people with atrial fibrillation (AF) is conflicting. This study aimed to examine the prevalence of frailty in hospitalised older patients with AF and its relationship with OAC prescription during admission. The secondary aim was to examine the association between frailty and rate/rhythm control medication prescriptions.

**Methods:** This retrospective observational study included adults aged ≥65 with AF admitted to six hospitals in Australia in 2022. Frailty was defined by a Frailty Index ≥0.25. Logistic regression models were applied to examine the association between frailty and the prescriptions of OAC, rate-control and rhythm-control drugs during hospitalisation. Results are presented as odds ratios and 95% confidence intervals (CI).

**Results:** There were 685 patients, with a mean age of 82.6(SD 8.3), 49.8% female and 42.8% identified as frail. Overall, 75.6% were prescribed OAC (67.9% in the frail versus 81.4% in the non-frail, p<0.001), 37.7% received rate-control drugs (42.0% in the frail versus 34.4% in the non-frail, p=0.044), 27.3% received rhythm-control drugs (22.9% in the frail versus 30.6% in the non-frail, p=0.024). The adjusted odds ratios of frailty on prescriptions were 0.58 (95% CI 0.39-0.86) for OAC, 1.75 (95%CI 1.22-2.52) for rate-control drugs, and 0.83 (95%CI 0.55-1.24) for rhythm-control drugs.

**Conclusions:** The study revealed a high prevalence of frailty in older inpatients with AF. Frailty was associated with reduced likelihood of prescription of OAC during admission and increased likelihood of prescribing rate-control medications, with no independent impact on rhythm-control therapy. Further studies are needed to understand these prescribing patterns.

## Introduction

Atrial fibrillation (AF) is the most prevalent arrhythmia globally and significantly contributes to stroke, posing an increasing burden on older people and healthcare systems.^1,2^ AF-related strokes are usually severe, resulting in chronic disability or death, and incur higher healthcare costs compared to non-AF strokes.^1^ The prevalence and incidence of AF increase significantly with age.^3,4^ It is predicted that between 2014 and 2034 the number of people with AF in Australia will double among older age groups (from 200,638 to 414,377 people with AF among those aged ≥75 years) and will increase 2.5-fold among men aged ≥85 (from 29,370 to 71,582).^5^ In Australia, more than half of all strokes occurred in patients aged 75 or older.^6,7^ AF was responsible for 36% of all ischemic strokes, of which 85% occurred in people with inadequate anticoagulation.^7^ Complications related to AF treatment also increase markedly in older adults, who face high risks of stroke, bleeding, and mortality.^1,2^

AF treatment focuses on stroke prevention through antithrombotic therapy, symptom reduction via rate-control or rhythm-control strategies, and managing associated medical conditions.^8^ Stroke prophylaxis with oral anticoagulation (OAC) is critical to AF management, and studies have shown that the benefits of direct OACs, compared with warfarin, extend to older adults.^1,2^ Furthermore, what constitutes appropriate anticoagulation, rate and rhythm control in older and frail patients is controversial.^1,9,10^ Digoxin may be useful for rate control, but associations with increased mortality limit its use.^10,11^ Syncope and fall-related injuries have been shown to be more common in older adults when OACs are used with antiarrhythmic drugs.^10,12^

As the population ages, the prevalence and clinical importance of frailty continue to rise.^13^ Frailty is a clinical syndrome resulting from multisystem impairments and characterised by increased vulnerability and disabilities.^14^ Multiple physiological factors are thought to be involved in the development of frailty, including the cardiovascular systems and thrombotic pathways.^15,16^ A relationship between frailty and cardiovascular disease has been observed, with frailty strongly linked to ischemic heart disease, heart failure and atrial fibrillation.^17,18^ Frailty predicts adverse outcomes such as loss of independence, increasing hospitalisations, and mortality in older patients, particularly those with cardiovascular diseases, and is associated with multimorbidity and polypharmacy.^17,19,20^ Clinical drug trials have traditionally excluded older adults with frailty due to concerns regarding their capacity to endure the treatments, and hence there is limited data from subgroup analyses by frailty status from randomised controlled trials on anticoagulants in older people.^21^ In older people, the lack of evidence from frail populations, concerns about medication adherence, and consideration of differences in pharmacokinetics, efficacy, and safety in the context of frailty, multimorbidity and polypharmacy may impact physicians’ decision to prescribe anticoagulants.^10,22^ Evidence of the impact of frailty on anticoagulant prescription in older people with AF is conflicting. Some studies suggested that OACs were underutilised in older people with frailty while others have not found this.^9,23,24^ The observed heterogeneity of the relationship between frailty and OAC prescriptions may be explained by the differences in study settings and frailty definitions used in these studies.

In this study, we aim to examine the association between frailty and OAC prescription in older hospitalised patients with AF during admission, taking into consideration the reasons for admission, services received, as well as other sociodemographic and clinical characteristics that may impact prescribing. The secondary aim was to examine the association between frailty and the prescription of rate/rhythm control medications during admission.

## Methods

### Study design

This is a retrospective observational cohort study, based on a dataset collected for another project (Optimising Quality Use of Medicines in Hospital to Improve Outcomes in Older People) (ACTRN12622000374763). The study was approved by the Northern Sydney Local Health District Human Research Ethics Committee (2023/ETH00422).

### Participants

Inclusion criteria: Adults aged ≥ 65 years admitted to six hospitals in the Northern Sydney Local Health District and Central Coast Local Health District, New South Wales, Australia between 1 June 2022 and 31 August 2022 with a diagnosis of AF (defined using the ICD-10- AM code) were included for this study. These included two tertiary referral hospitals, two major metropolitan hospitals, one district general metropolitan hospital and one subacute rehabilitation hospital. When a patient was admitted to hospitals more than once during the study period, we included their first admission which had a length of hospitalisation >48 hours. Exclusion criteria: Admissions with length of stay <48 hours were excluded from this study.

### Variable definition

Frailty: Frailty is the predictive variable of interest for the prescriptions of OAC, rate control and rhythm control medications. Frailty was defined by a Frailty Index constructed using information from the electronic medical records including the comprehensive care assessment, with a reference to the recent published work by Lo, Fujita and colleagues on the development and validation of a Frailty Index based on data routinely collected across multiple domains in NSW hospitals (electronic Frailty Index for Acute Hospitals, eFI- AH).^25,26^ The Frailty Index is one convenient method of quantifying frailty and has high predictive ability for adverse health outcomes in older people.^27^ The Frailty Index is based on the conceptualisation of frailty as an accumulation of deficits throughout life. It is constructed as the proportion of deficits present in an individual out of the total number of age-related health variables considered, with a value obtained from 0 to 1. While the Frailty Index is a continuous variable, for analyses in this study, the previously validated cut-point of 0.25 was applied to define frailty.^28,29^

Information on prescribed medications were extracted from the electronic medical record system. The prescribed medications were grouped into OAC (warfarin, dabigatran, apixaban, rivaroxaban), rate control drugs (digoxin, diltiazem, verapamil), and rhythm control drugs (amiodarone, flecainide, sotalol). A medication was considered prescribed if it was noted at any stage during the admission. However, it was not possible to determine if prescriptions were initiated or ceased during the admission.

### Covariates

The covariates included age, sex, Charlson Comorbidity Index, hospitals and services, and reasons for admission. The reasons for admission were obtained from notes in the medical records and were categorised into 18 groups based on conditions that could affect the prescription of the drugs of interest and their prevalence in the study cohort: chest pain, shortness of breath, generally unwell, dizziness, altered level of consciousness, stroke/ transient ischemic attack (TIA)/ embolism, heart failure, myocardial infection, rhythm disorder, infection, surgery/procedure, falls, injury/fracture, gastrointestinal disorder, musculoskeletal pain, bleeding, acute kidney injury, and other.

### Sample size justification

For this study, we estimated that at least 500 patients with AF are needed to detect a difference in OAC prescription between the frail and the non-frail (with a power of 90% and alpha=0.05). This power analysis is based on our previous study at Royal North Shore Hospital during the period from 2012 to 2014, in which the prescription rate of anticoagulants was 49% in the frail and 63% in the non-frail patients with AF. ^24^

### Statistical analysis

Continuous variables are presented as mean ± standard deviation (SD) or median with interquartile range, and categorical variables as frequency and percentage. Comparisons between frail and non-frail participants were assessed using the Chi-square test or Fisher’s exact test for categorical variables and Student’s t-test or Mann-Whitney test for continuous variables.

To examine the association of frailty with the prescriptions of OAC, rate and rhythm control drugs in older patients with AF, logistic regression models were applied with frailty as the explanatory variable of interest, adjusted for the covariates mentioned above. Results are presented as odds ratios (OR) and 95% confidence intervals. Two-tailed P values < 0.05 are considered significant. Analysis of the data was performed using Python, SPSS and/or R.

## Results

A total of 685 patients were included in this analysis. They had a mean age of 82.6 (± 8.3) years, and 49.8% were female. Figure 1 presents the study flowchart.

**Figure 1.**
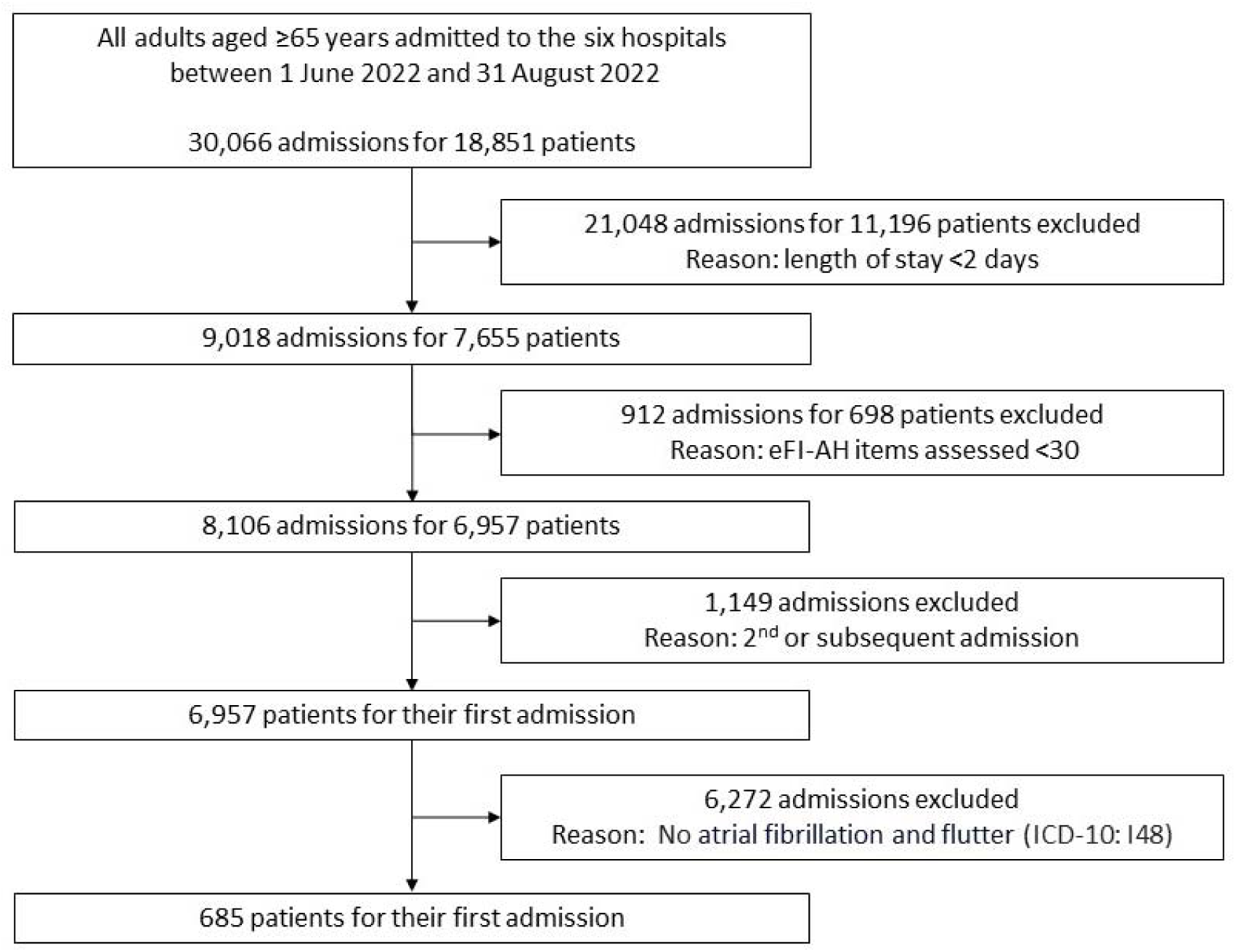
The Study Flowchart.

**Table 1.** Patient characteristics by frailty status.

| Characteristics | All<br>(N=685) | Non-frail<br>(N=392) | Frail<br>(N=293) | P |
| --- | --- | --- | --- | --- |
| Age | 82.6 ± 8.33 | 81.2 ± 8.15 | 84.5 ± 8.20 | <0.001 |
| Female | 341 (49.8%) | 186 (47.4%) | 155 (52.9%) | 0.158 |
| Language |  |  |  |  |
| English speaking | 636 (92.8%) | 374 (95.4%) | 262 (89.4%) | 0.003 |
| Non-English speaking | 49 (7.2%) | 18 (4.6%) | 31 (10.6%) |  |
| Marital status |  |  |  |  |
| Married | 335 (48.9%) | 213 (54.3%) | 122 (41.6%) | 0.019 |
| Widowed | 210 (30.7%) | 106 (27.0%) | 104 (35.5%) |  |
| Divorced/Separated | 82 (12.0%) | 41 (10.5%) | 41 (14.0%) |  |
| Never married | 47 (6.9%) | 27 (6.9%) | 20 (6.8%) |  |
| Unknown | 11 (1.6%) | 5 (1.3%) | 6 (2.0%) |  |
| Smoking |  |  |  |  |
| Never smoked | 311 (45.4%) | 191 (48.7%) | 120 (41.0%) | 0.043 |
| Smokers/ex-smokers | 374 (54.6%) | 201 (51.3%) | 173 (59.0%) |  |
| Charlson Comorbidity Index | 1.49 ± 1.62 | 1.07 ± 1.29 | 2.05 ± 1.83 | <0.001 |
| Services |  |  |  |  |
| Cardiology | 204 (29.8%) | 155 (39.5%) | 49 (16.7%) | <0.001 |
| General Medicine | 134 (19.6%) | 81 (20.7%) | 53 (18.1%) |  |
| Surgery | 106 (15.5%) | 53 (13.5%) | 53 (18.1%) |  |
| Neurology | 83 (12.1%) | 41 (10.5%) | 42 (14.3%) |  |
| Geriatric Medicine | 75 (10.9%) | 23 (5.9%) | 52 (17.7%) |  |
| Other | 83 (12.1%) | 39 (9.9%) | 44 (15.0%) |  |

Using the Frailty Index cut-off point of 0.25, 42.8% of the patients in the sample were identified as being frail. The distribution of the Frailty Index (eFI-AH) in the sample is presented in Appendix 1.

Compared to non-frail patients, frail patients were older (mean age 84.5 ± 8.2 in the frail versus 81.2 ± 8.2 in the non-frail, p <0.001). The frail group had higher proportion of people from a non-English speaking background (10.6% compared to 4.6% in the non-frail, p = 0.003) and a higher proportion of smoking (59.0% compared to 51.3% in the non-frail, p = 0.043). They also had higher mean Charlson Comorbidity Index value (2.1 ± 1.8 compared to 1.1 ± 1.3 in the non-frail, p <0.001).

### Reasons for admission

The most common reasons for admission included infection, shortness of breath, stroke/TIA/embolism, and falls. Frail patients were more likely to present to hospitals due to falls, fracture/injury, while non-frail patients were more likely to present with cardiovascular-related reasons such as heart failure, chest pain, myocardial infarction, rhythm disorders, and dizziness. (Figure 2) Most patients with AF were admitted to Cardiology services (29.8%), followed by General Medicine (19.6%), Surgery (15.5%), Neurology (12.1%), Geriatric Medicine (10.9%), and other services (12.1%).

**Figure 2.**
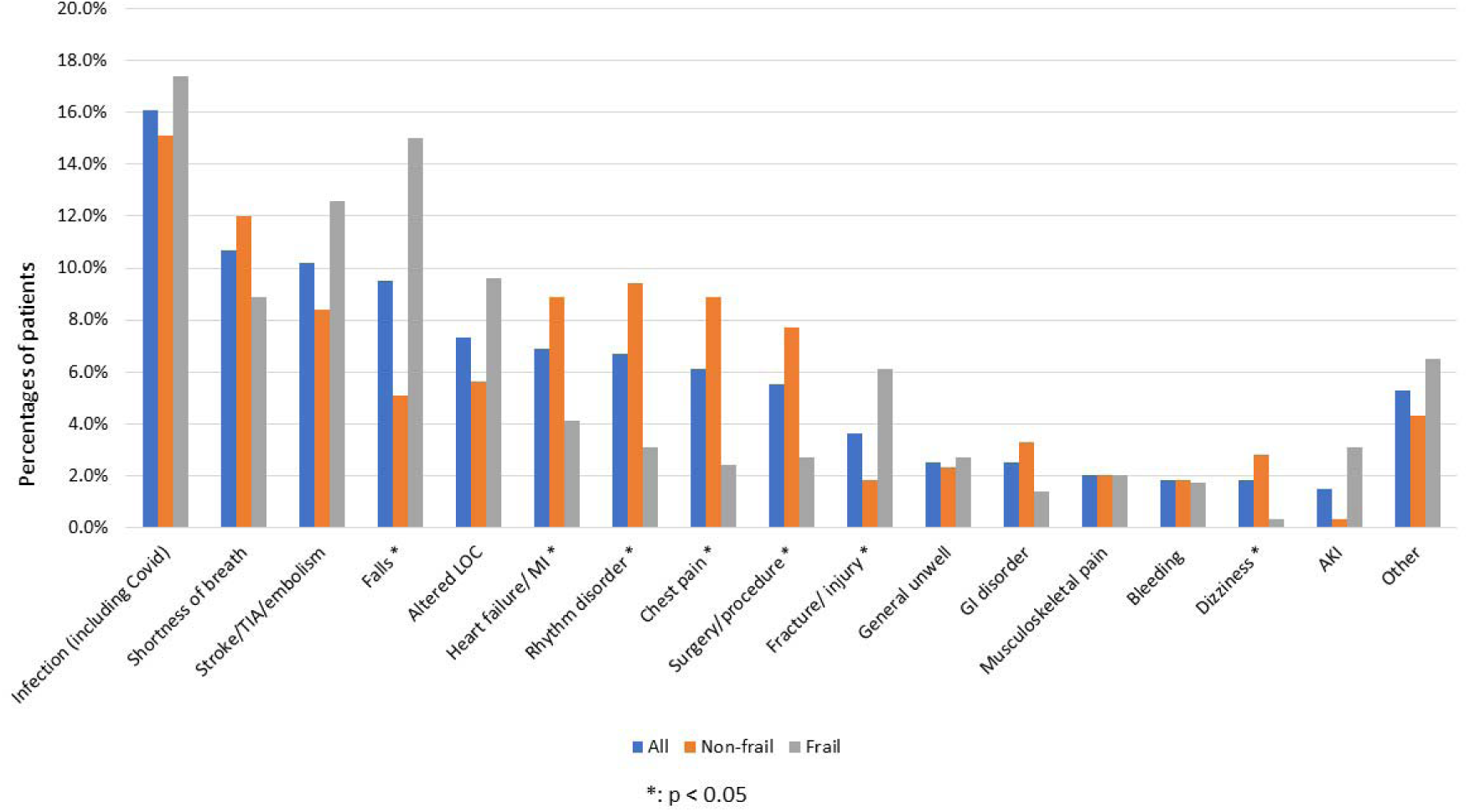
Reasons for admission among 685 patients with atrial fibrillation. TIA: transient ischaemic attack; LOC: level of consciousness; MI: myocardial infarction; GI: gastrointestinal; AKI: acute kidney injury.

### Prescription of oral anticoagulants and predictors

Overall, 75.6% of the patients were prescribed OACs (67.9% in the frail compared to 81.4% in the non-frail, p <0.001). The adjusted odds ratio of frailty on OAC prescriptions during admission was 0.58 (95% CI 0.39 – 0.86), adjusted for age, sex, the Charlson Comorbidity Index, reasons for admission, hospitals and services received (Figure 4). Other factors that were independently associated with OAC prescriptions included presenting with falls (adjusted OR 0.53, 95%CI 0.29 – 0.99), presenting with gastrointestinal disorders (adjusted OR 0.28, 95%CI 0.10 – 0.79), and admitted to a Cardiology Service (adjusted OR 2.86, 95%CI 1.66 – 4.92).

**Figure 3.**
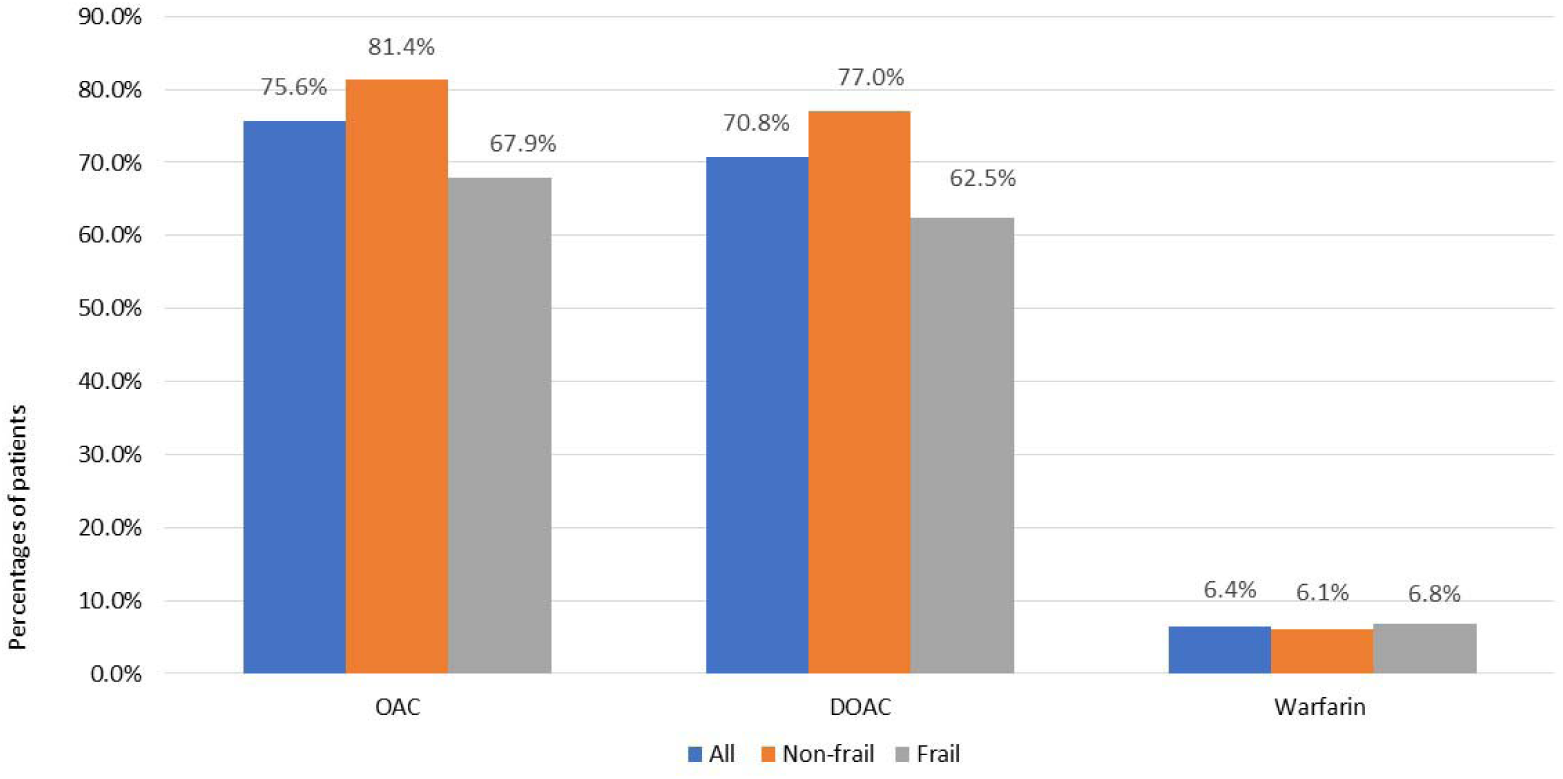
Percentages of patients prescribed oral anticoagulants during admission among 685 patients with atrial fibrillation, stratified by frailty. OAC: oral anticoagulant; DOAC: direct oral anticoagulant.

**Figure 4.**
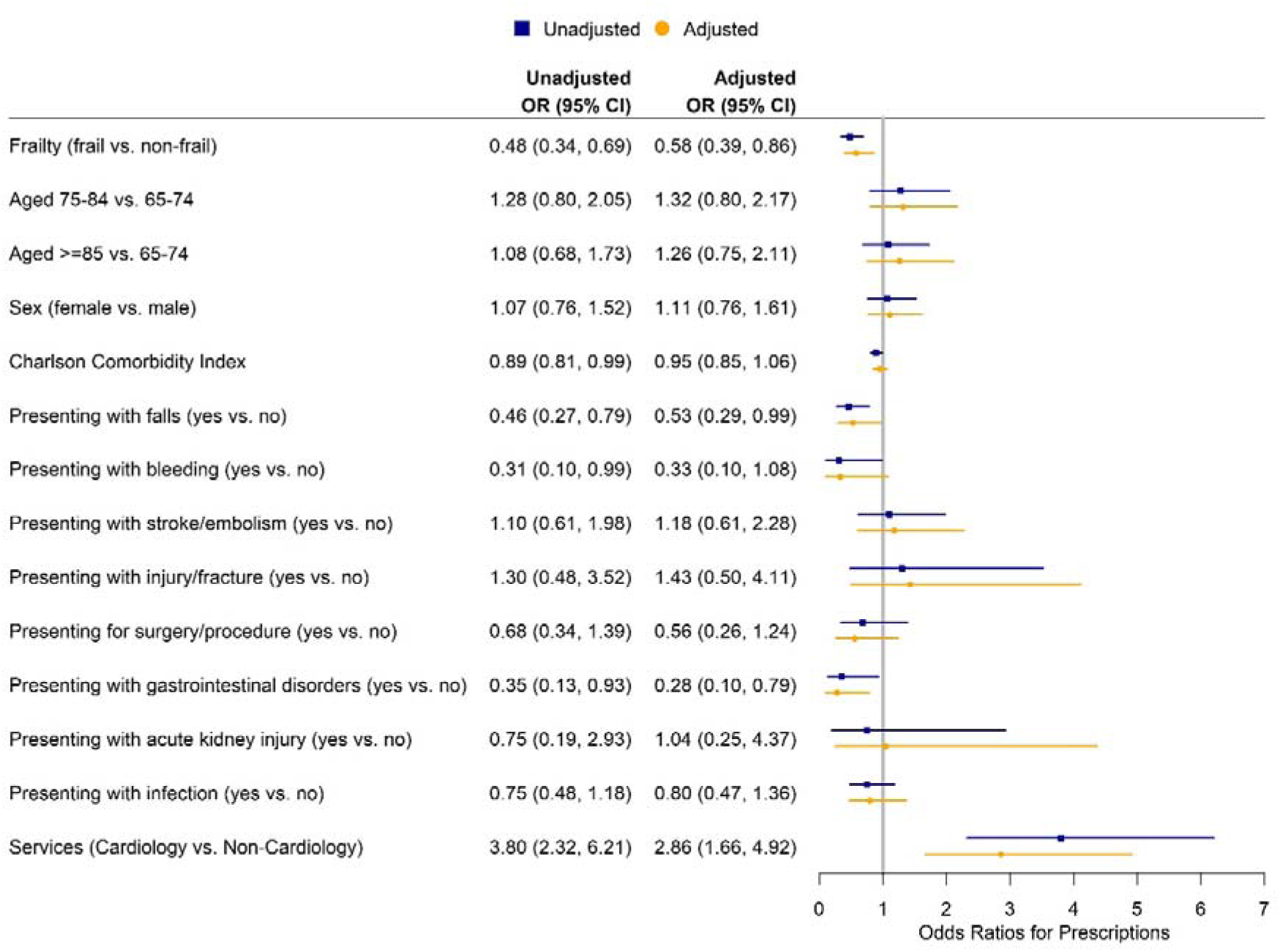
Factors associated with prescriptions of oral anticoagulants.

### Prescriptions of rate and rhythm control drugs

Overall, 37.7% of the participants received rate control drugs (42.0% in the frail versus 34.4% in the non-frail, p = 0.044), while 27.3% received rhythm control drugs (22.9% in the frail compared to 30.6% in the non-frail, p = 0.024). A significant proportion of patients (47%) were not prescribed any of these medications (Appendix 3).

On the adjusted model, frailty (adjusted OR 1.75, 95% CI 1.22 – 2.52) and being admitted to a Cardiology service (adjusted OR 1.53, 95%CI 1.02 – 2.30) significantly increased the odds of being prescribed rate control drugs during admission. (Appendix 4)

Being admitted to a Cardiology service was also associated with an increased odds of being prescribed rhythm control drugs (adjusted OR 2.56, 95%CI 1.65 – 3.97). Older age was associated with reduced odds of being prescribed these medications (adjusted OR 0.63, 95%CI 0.40 – 1.00 for aged 75 – 85 vs. aged 65 – 74, and 0.54 (0.33 – 0.88) for aged ≥ 85 vs. aged 65 – 74). Frailty had no independent relationship with rhythm control drug prescriptions (adjusted OR 0.83, 95% CI 0.55 – 1.24). (Appendix 5)

## Discussion

In a study involving 685 older hospitalised patients with AF, it was found that there was a high prevalence of frailty in the studied cohort. The presence of frailty decreased the odds of being prescribed anticoagulants during admission by half, even after adjusting for factors such as age, sex, reasons for admission, comorbidities, and hospital service. Frailty did not affect the prescription of rhythm control medications but was linked to an increased odds of receiving rate control therapy during admission.

Atrial fibrillation is very common in older people. The presence of frailty in an older person with AF can make it challenging for clinicians to determine the appropriate dosage and monitoring for anticoagulant therapy. Frailty may also affect a patient’s ability to adhere to their anticoagulant regimen, which can increase the risk of stroke, bleeding and other complications associated with anticoagulants.^30^ However, current guidelines provide limited specific guidance for the treatment of this condition in frail patients.^8,31^ There is conflicting evidence on the impact of frailty on the prescription of OAC in patients with AF. Some studies suggest that frailty and geriatric syndromes are associated with non-prescription of anticoagulants ^32,33^ while others have not found this.^34,35^ A systematic review and meta-analysis conducted by Wilkinson and colleagues in 2019 showed that frailty was associated with decreased OAC prescription at hospital admission (pooled adjusted OR 0.45, 95%CI 0.22–0.93).^9^ However, they found that frailty had no statistically significant association with OAC prescription at discharge (pooled adjusted OR 0.40, 95% CI 0.13−1.23).^9^ Recently, Bul and colleagues conducted a systematic review on frailty and oral anticoagulant prescription in adults with AF and found that only 52% of the frail population were anticoagulated compared to 67% in the non-frail.^23^

Compared with other local studies conducted in the past three decades in one of the six current study hospitals, we observed that the prevalence of anticoagulant prescription in older patients with AF has increased over time: from 35.0% in 1997 ^36^ to 39.1% in 2007 (23.6% in the frail and 66.3% in the non-frail) ^33^ and 55.7% in 2014 (49.3% in the frail, 62.6% in the non-frail). The increase in anticoagulation in older patients with AF, including the frail, over this period may reflect the uptake of novel/direct oral anticoagulants and the translation of new evidence into clinical practice.^37^

The increased odds of people with frailty being prescribed rate control therapy (digoxin, diltiazem or verapamil) during admission was consistent with our previous observation for digoxin at one of the study hospitals but remains difficult to explain.^24^ Digoxin, the commonest rate control therapy prescribed in our cohort, has a narrow therapeutic index, is renally cleared and frailty independently impairs renal function.^38^ Digoxin is considered potentially inappropriate for first line rate control treatment of AF by the American Geriatrics Society Beers criteria.^39^ Digoxin is a major cause of hospital admissions for adverse drug reactions in geriatric patients.^40^ To our knowledge, this has not been described previously and highlights the importance of comprehensive medication review for frail older adults.

## Strengths and limitations

This was a multi-centred retrospective observational study and conducted based on high quality hospital data and comprised a large sample of very old and frail hospitalised patients who were admitted to metropolitan and regional hospitals of different sizes and acuity. Frailty was defined based on a Frailty Index using data from the routine electronic medical records, the eFI-AH, which has been validated in previous studies.^25,26^

A major limitation of this study is that it was conducted in public hospital settings in NSW only, which may not be nationally generalisable. We were unable to capture details such as the duration of AF and types of AF. Therefore, the study findings should be cautiously interpreted and extended to all older people with AF. We were unable to determine changes to prescribing made in hospital.

## Conclusion

Our study found a high prevalence of frailty in older inpatients with AF. Frailty was associated with reduced likelihood of receiving anticoagulants during admission. Further studies are needed to understand the reasons for lower prevalence of anti-coagulation in frail patients with AF. It is crucial for healthcare providers to routinely assess and monitor frailty and to regularly review prescribing in older patients with AF to individualise therapy in this population, who are at high risk of complications of AF and of severe adverse drug reactions.

## Data Availability

All data produced in the present study are available upon reasonable request to the authors.

## Acknowledgement

TNN was supported by the Sydney Health Partners Geriatric Medicine Clinical Academic Group Early Career Researcher’s Award 2022. KF is supported by a grant from the Australian National Health and Medical Research Council Targeted Call for Research into Frailty in Hospital Care (APP 1174447).

## Author Contributions

TNN and SH conceptualised the study. TNN conducted the literature search, led the analysis, and drafted the manuscript. All authors (TNN, KJ and SH) contributed to the design of the study, interpreted the results, and critically revised the manuscript. All authors accept responsibility to submit for publication and gave approval for the final version to be published.

## Appendices

## Appendix 1. The distribution of the Frailty Index in the study cohort

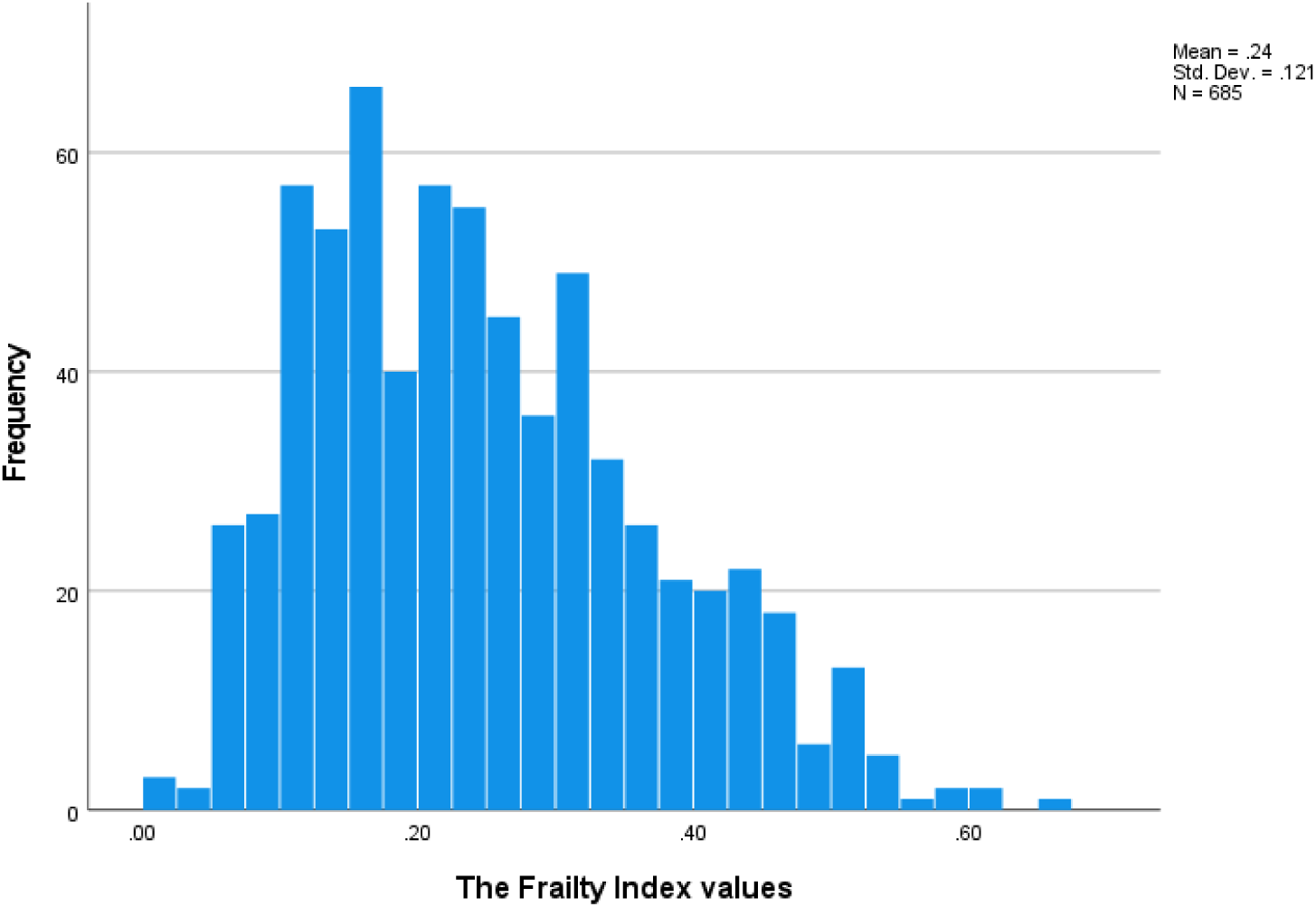

Frailty Index was calculated using the electronic Frailty Index for Acute Hospitals (eFI-AH)

## Appendix 2. Prescriptions of rate and rhythm control drugs

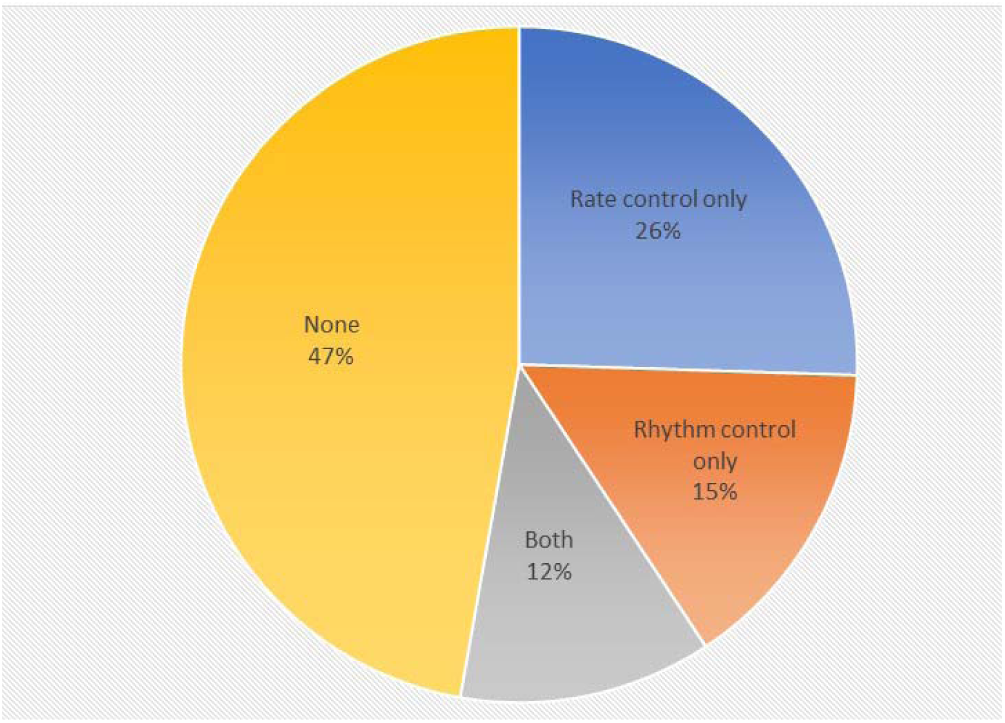

## Appendix 3. Prevalence of prescriptions of rate and rhythm control drugs by frailty status

|  | **All**<br>**(N=685)** | **Non-frail**<br>**(N=392)** | **Frail**<br>**(N=293)** | ***p*-value** |
| --- | --- | --- | --- | --- |
| Rate control | 258 (37.7%) | **135 (34.4%)** | **123 (42.0%)** | **0.044** |
| Digoxin | 238 (34.7%) | 121 (30.9%) | 117 (39.9%) | 0.014 |
| Diltiazem | 28 (4.1%) | 19 (4.8%) | 9 (3.1%) | 0.246 |
| Verapamil | 18 (2.6%) | 9 (2.3%) | 9 (3.1%) | 0.530 |
| Rhythm control | 187 (27.3%) | **120 (30.6%)** | **67 (22.9%)** | **0.024** |
| Amiodarone | 111 (16.2%) | 68 (17.3%) | 43 (14.7%) | 0.348 |
| Flecainide | 26 (3.8%) | 16 (4.1%) | 10 (3.4%) | 0.650 |
| Sotalol | 64 (9.3%) | 45 (11.5%) | 19 (6.5%) | 0.026 |

## Appendix 4. Factors associated with rate control therapies

| Variables | Univariable analysis |  | Multivariable analysis |  |
| --- | --- | --- | --- | --- |
|  | Unadjusted OR for prescription of rate control drugs (95%CI) | *p*-value | Adjusted OR for prescription of rate control drugs (95%CI) | *p*-value |
| Frailty |  |  |  |  |
| Non-frail (reference) | 1 |  | 1 |  |
| Frail | 1.38 (1.01 – 1.88) | 0.044 | 1.75 (1.22 – 2.52) | 0.002 |
| Age group |  | 0.927 |  | 0.912 |
| 65-74 (reference) | 1 |  | 1 |  |
| 75-84 | 1.08 (0.70 – 1.65) | 0.733 | 1.09 (0.70 – 1.69) | 0.704 <sup>a</sup> |
| ≥ 85 | 1.08 (0.71 – 1.66) | 0.712 | 1.03 (0.65 – 1.62) | 0.915 <sup>b</sup> |
| Sex |  |  |  |  |
| Male (reference) | 1 |  | 1 |  |
| Female | 1.40 (1.03 – 1.91) | 0.033 | 1.36 (0.98 – 1.87) | 0.066 |
| Charlson Comorbidity Index | 1.00 (0.91 – 1.10) | 0.992 | 0.98 (0.88 – 1.09) | 0.705 |
| Presenting with falls | 0.68 (0.39 – 1.18) | 0.169 | 0.61 (0.33 – 1.15) | 0.126 |
| Presenting with injury/fracture | 1.11 (0.49 – 2.50) | 0.806 | 0.97 (0.41 – 2.29) | 0.935 |
| Presenting with rhythm disorder | 2.28 (1.25 – 4.18) | 0.008 | 1.78 (0.92 – 3.44) | 0.087 |
| Presenting with bleeding | 0.33 (0.07 – 1.50) | 0.150 | 0.34 (0.07 – 1.63) | 0.177 |
| Presenting with stroke/embolism | 0.63 (0.37 – 1.09) | 0.100 | 0.65 (0.36 – 1.20) | 0.169 |
| Presenting for surgery/procedure | 0.66 (0.32 – 1.35) | 0.257 | 0.78 (0.36 – 1.68) | 0.519 |
| Presenting with gastrointestinal disorders | 1.16 (0.44 – 3.09) | 0.762 | 1.27 (0.46 – 3.53) | 0.646 |
| Presenting with acute kidney injury | 0.41 (0.09 – 1.94) | 0.261 | 0.35 (0.07 – 1.71) | 0.194 |
| Presenting with infection | 1.18 (0.78 – 1.78) | 0.444 | 1.17 (0.72 – 1.89) | 0.523 |
| Services |  |  |  |  |
| Non-Cardiology (reference) | 1 |  | 1 |  |
| Cardiology | 1.61 (1.15 – 2.24) | 0.005 | 1.53 (1.02 – 2.30) | 0.042 |

<sup>a</sup> *p*-value for comparison between the age group 75-84 versus age group 65-74; <sup>b</sup> *p*-value for comparison between the age group ≥85 versus age group 65-74

## Appendix 5. Factors associated with rhythm control therapies

| Variables | Univariable analysis | Multivariable analysis |
| --- | --- | --- |

|  | Unadjusted OR for<br>prescription of rhythm<br>control drugs (95%CI) | *p*-value | Adjusted OR for<br>prescription of rhythm<br>control drugs (95%CI) | *p*-value |
| --- | --- | --- | --- | --- |
| Frailty |  |  |  |  |
| Non-frail (reference) | 1 |  | 1 |  |
| Frail | 0.67 (0.48 – 0.95) | 0.025 | 0.83 (0.55 – 1.24) | 0.353 |
| Age group |  | 0.012 |  | 0.040 |
| 65-74 (reference) | 1 |  | 1 |  |
| 75-84 | 0.62 (0.40 – 0.96) | 0.032 | 0.63 (0.40 – 1.00) | 0.050<sup>a</sup> |
| ≥ 85 | 0.51 (0.33 – 0.80) | 0.003 | 0.54 (0.33 – 0.88) | 0.013<sup>b</sup> |
| Sex |  |  |  |  |
| Female (reference) | 1 | 1 | 1 |  |
| Female | 0.91 (0.65 – 1.28) | 0.596 | 1.02 (0.71 – 1.46) | 0.920 |
| Charlson Comorbidity<br>Index | 1.01 (0.91 – 1.12) | 0.881 | 1.10 (0.98 – 1.23) | 0.126 |
| Presenting with falls | 0.96 (0.54 – 1.72) | 0.889 | 1.51 (0.78 – 2.93) | 0.222 |
| Presenting with<br>injury/fracture | 1.04 (0.43 – 2.53) | 0.936 | 1.57 (0.60 – 4.10) | 0.354 |
| Presenting with rhythm<br>disorder | 2.63 (1.44 – 4.82) | 0.002 | 1.49 (0.77 – 2.92) | 0.239 |
| Presenting with bleeding | 0.24 (0.03 – 1.86) | 0.171 | 0.36 (0.05 – 2.90) | 0.338 |
| Presenting with<br>stroke/embolism | 0.41 (0.21 – 0.82) | 0.012 | 0.69 (0.32 – 1.46) | 0.328 |
| Presenting for<br>surgery/procedure | 1.41 (0.71 – 2.83) | 0.327 | 1.45 (0.68 – 3.11) | 0.338 |
| Presenting with<br>gastrointestinal disorders | 1.47 (0.54 – 4.03) | 0.456 | 2.09 (0.71 – 6.10) | 0.179 |
| Presenting with acute<br>kidney injury | 1.14 (0.29 – 4.47) | 0.847 | 1.55 (0.37 – 6.44) | 0.544 |
| Presenting with infection | 0.66 (0.40 – 1.09) | 0.102 | 0.93 (0.53 – 1.65) | 0.811 |
| Services |  |  |  |  |
| Non-Cardiology<br>(reference) | 1 |  | 1 |  |
| Cardiology | 2.74 (1.92 – 3.91) | <0.001 | 2.56 (1.65 – 3.97) | <0.001 |

<sup>a</sup> p-value for comparison between the age group 75-84 versus age group 65-74; <sup>b</sup> p-value for comparison between the age group $\geq 85$ versus age group 65-74

